# Multi-model forecasts of the ongoing Ebola epidemic in the Democratic Republic of Congo, March – October 2019

**DOI:** 10.1101/2020.06.07.20124867

**Authors:** Kimberlyn Roosa, Amna Tariq, Ping Yan, James M. Hyman, Gerardo Chowell

## Abstract

The 2018-20 Ebola outbreak in the Democratic Republic of the Congo is the first to occur in an armed conflict zone. The resulting impact on population movement, treatment centers, and surveillance has created an unprecedented challenge for real-time epidemic forecasting. Most standard mathematical models cannot capture the observed incidence trajectory when it deviates from a traditional epidemic logistic curve. We fit seven dynamic models of increasing complexity to the incidence data published in the World Health Organization Situation Reports, after adjusting for reporting delays. These models include a simple logistic model, a Richards model, an endemic Richards model, a double logistic growth model, a multi-model approach, and two sub-epidemic models. We analyze model fit to the data and compare real-time forecasts throughout the ongoing epidemic across 29 weeks from March 11 to September 23, 2019. We observe that the modest extensions presented allow for capturing a wide range of epidemic behavior. The multi-model approach yields the most reliable forecasts on average for this application, and the presented extensions improve model flexibility and forecasting accuracy, even in the context of limited epidemiological data.

## Introduction

There is a long, rich history of using mathematical models to study the spread and control of infectious diseases (1-3). For instance, mathematical models can provide insight on the impact of different transmission mechanisms and interventions (4-6), estimate transmission potential across different pathogens and social settings (7, 8), and evaluate optimal strategies for resource allocation (9, 10). Mathematical models can forecast, identify, and predict the morbidity and mortality patterns in infectious disease outbreaks in near real time (e.g., (10, 11)). Public health officials can use the model short-term projections to inform public health interventions during an outbreak (4, 12-18).

Many modeling studies rely on historical epidemic data to evaluate the effectiveness of the model for forecasting an epidemic (5, 6, 13, 15). In contrast, real-time studies aim to generate predictions as the epidemic unfolds (4, 7, 10-12, 19-21). These real-time studies present with additional challenges, as surveillance data is often affected by underreporting, misclassification, and reporting delays (21, 22). Fortunately, standard statistical methods can be useful to adjust short-term incidence trends for reporting delays and “nowcast” data in real time (21, 23, 24).

The 2018-20 Ebola epidemic in the Democratic Republic of the Congo (DRC) was initially declared on August 1, 2018. As of April 26, 2020, a total of 3461 cases have been reported, mostly in the provinces of North Kivu and Ituri, (with 6 cases from the province of South Kivu) (25). The outbreak has now largely been brought under control; however, small resurgences are still being reported over a year and a half after the start of the outbreak. Despite vaccination and other preventative efforts, the outbreak has persisted largely due to long-standing conflict in the region, including recurrent violent attacks targeting Ebola treatment centers and healthcare teams (25-27). Particularly, regions of North Kivu and Ituri have been destabilized, leading to conflict from more than 70 armed militant groups (28). In addition to violence, a complicated history of humanitarian intervention has hindered the Ebola response efforts, impacting epidemiological surveillance and contact tracing efforts, including temporary suspension of Ebola response activities (22, 26, 28, 29). The multiple Ebola resurgences associated with these instabilities have resulted in a multimodal incidence pattern (see Figure S1) (7, 30). The complex characteristics and trajectory of this outbreak pose an unprecedented challenge for forecasting the trajectory of the epidemic in real time.

In February 2019, a sharp increase in cases and transmission were observed, coinciding with deteriorating security, targeted attacks on response teams, and decreasing trust in the Ebola response efforts (31, 32). Previous studies have provided real-time forecasts at different time points of the 2018-20 Ebola epidemic in the DRC (Figure S1) using various approaches, including a semi-structured model that relies on nowcasting (21), stochastic and auto-regressive models that incorporate historical data (20), as well as a sub-epidemic wave framework (30), which we also use here. While each of these approaches performed well for fitting and forecasting the trajectory of the outbreak in 2018 and early 2019, each model failed to predict the case resurgence observed in February 2019, resulting in forecasts that drastically underestimated the true cumulative case count to date. Therefore, we focus model calibration in this study on the large 2019 resurgence to better project the upcoming epidemic trajectory. This also allows for the implementation of simpler models, including models that only allow for a single peak.

We systematically compare real-time forecasts (1 - 4 weeks ahead) for the ongoing Ebola epidemic in the DRC using seven dynamic models of variable complexity. Our models range from simple scalar differential equation models, such as the standard logistic growth and Richards models, to more complex dynamic models that capture a diversity of epidemic trajectories, such as multimodal outbreaks. These include extensions of the recently developed sub-epidemic wave framework consisting of systems of differential equations (30), an extended Richards model that incorporates an endemic state, and a double logistic growth model that supports incidence curves with two peaks (33). We also present a performance-based multi-model approach that incorporates the four single equation models in order of increasing complexity. We stratify forecasting performance within specific forecasting phases, as defined by the multi-model approach.

## Data and Methods

### Incidence data of the DRC Ebola epidemic and adjusting for reporting delays

We retrieve weekly case incidence data for the 2018-20 Ebola epidemic in the DRC from the epidemic curves published weekly in the World Health Organization (WHO) Situation Reports (25). The recurrent violent attacks and widespread public distrust have hindered the Ebola surveillance and containment efforts (26, 34) and resulted in delays in reporting the true incidence curve (22). Outbreak curves describing epidemic spread in near real-time can be distorted by reporting delays, so we adjust the crude incidence for reporting delays using statistical methods.

Reporting delay is defined as the time lag between the time of onset and the time when the case is reported and entered into the database (33). Reporting delays occur for multiple reasons including difficulty in tracing and monitoring cases, attacks on health workers and health centers, resistance of sick individuals to seek treatment as soon as symptoms occur, inefficient surveillance systems, and population mobility (35). We use a non-parametric actuaries method that adapts survival analysis for use with right truncated data by employing point estimation based on reverse time hazards to statistically adjust for reporting delays based on the empirical distribution of the delays (23, 36-38). This allows us to estimate the number of occurred but not yet reported events at a given point in time due to incomplete case reporting.

This method involves expressing the conditional reporting delay distribution as the product of conditional probabilities. The adjusted incidence data are obtained by appropriately dividing the observed number of cases by the reporting delay distribution. Then we derive the 95% prediction limits using the statistical estimations introduced by Lawless, modified to assume non-stationary reporting delay probabilities, considering the most recent reporting period. This yields robust and realistic 95% prediction intervals (38, 39). We use weekly time intervals as a compromise between maximizing the temporal resolution and reporting irregularities in batch reporting of case counts and inaccuracies in retrospectively ascertained dates of onset. We sequentially analyze incidence data from Situation Reports as more information becomes available to adjust for reporting delays. All data are being made publicly available in a repository (40).

### Model calibration and forecasting approach

We conducted 29 week-to-week forecasts between March 11 and September 23, 2019. Each forecast was fit to the reporting delay adjusted weekly incidence from data reported in Situation Reports 33 – 61, between March 19, 2019, and October 1, 2019. The uncertainty in the reporting delay is greatest in the most recently reported (last observed) weekly incidence data point; thus, we exclude the last weekly incidence data point in the analysis (lag of 1 week) (Figure S9). The first model calibration process relies on five incidence weeks: February 11 – March 11, 2019, with the latest snapshot of the epidemic corresponding to Situation Report 33 (March 19, 2019). Sequentially, models are re-calibrated each week using the most up-to-date adjusted incidence curve; meaning, the length of the calibration period increases by one week with each new weekly published WHO Situation Report (25).

We estimate the model parameters, Θ = (θ_1_, θ_2_, …, θ_m_), in Table S1 from the data using nonlinear least squares fitting to minimize the sum of squared errors between the model prediction *f*(*t, Θ*) and the data *y*_*t*_. The parameters 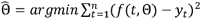 define the best - fit model 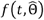. To test the uniqueness of the best-fit model, we initialize the parameters for the nonlinear least squares method over a wide range of feasible parameters from a uniform distribution using Latin hypercube sampling. Further, we fix the initial condition according to the first data point.

We use a parametric bootstrap approach to quantify parameter uncertainty and estimate prediction intervals, which involves resampling with replacement of incidence data assuming a Poisson error structure (41). Our calibration results represent *M =* 300 resampled data sets that are refit to obtain *M* new parameter estimates. Model fits are used to obtain 95% confidence intervals for each parameter (41).

Each of the *M* model fits is used to generate *m* = 30 simulated data curves with Poisson noise; these 9,000 (*M × m*) curves are then used to construct the 95% prediction intervals for the forecasting period of 1 – 4 weeks (*h* = 1, 2, 3, 4). We give a detailed description of this parameter estimation method in prior studies (41-43).

### Performance metrics

We used the following model performance metrics to assess the quality of the model fit and forecasting performance (*h =* 1 – 4 weeks ahead). For calibration performance, we compare model fit to the adjusted observed data; whereas, we compare forecasts with the raw incidence data reported four weeks ahead of the last date of the calibration period.

The mean squared error (MSE) and the mean absolute error (MAE) assess average deviations of the model to the observed data:

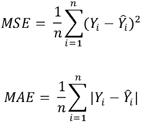

where *Y*_*i*_ is the data, 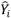 is the model prediction, and *n* is the number of data points in the interval. For the calibration period, *n* equals the number of data points calibrated to, and for the forecasting period, *n = h =* 1, 2, 3, 4 for 1 – 4 weeks ahead forecasts, respectively.

To assess model uncertainty and performance of prediction intervals, we use prediction interval coverage (PI coverage) and mean interval score (MIS) (44). Prediction interval coverage is the fraction of data points that fall within the 95% prediction interval; calculated as:

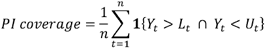

where *n* is the length of the period, *L*_*t*_ and *U*_*t*_ are the lower and upper bounds of the 95% prediction intervals, respectively, *Y*_*t*_ are the data, and **1** is an indicator variable that equals 1 if *Y*_*t*_ is in the specified interval and 0 otherwise.

The mean interval score considers the width of the interval as well as the coverage, with a penalty for data points not included within the prediction intervals. The MIS is calculated as

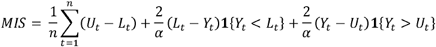

where *n* is the length of the period, *L*_*t*_ and *U*_*t*_ are the lower and upper bounds of the 95% prediction intervals, *Y*_*t*_ are the data, α is the significance level (α = 0.05), and **1** is an indicator variable that equals 1 if *Y*_*t*_ is in the specified interval and 0 otherwise (44). Therefore, if the PI coverage is 1, the MIS is the average width of the interval across each time point. For two models with equivalent PI coverage, a lower MIS indicates narrower intervals.

### Forecasting strategy

We evaluate short-term forecasts in real-time using seven dynamic models: four single-equation models of increasing complexity, a multi-model approach, and two sub-epidemic wave models whose complexity depends on the temporal pattern of the epidemic. Features such as number of parameters, number of equations, and ability to capture varying dynamics are provided in Table 1. A brief overview of the models is provided below, and the Supplement contains additional details to fully define the models.

**Table 1.** Real-time forecasting comparison of seven dynamic models. The multi-model approach encompasses the four single-equation models in the first four rows (Figure 1).

|  | # of fitting<br>parameters | # of<br>differential<br>equations | Supports<br>endemic state | Supports<br>two peaks | Supports<br>oscillations |
| --- | --- | --- | --- | --- | --- |
| **Logistic** | 2 | 1 | No | No | No |
| **Richards** | 3 | 1 | No | No | No |
| **Endemic**<br>**Richards** | 5 | 1 | Yes | No | No |
| **Double logistic** | 6 | 1 | Yes | Yes | No |
| **Multi-model** | 2 – 6 | 1 | Yes | Yes | No |
| **Sub-epidemic I** | 5 | $\geq 1$ (*n*) | Yes | Yes | Yes |
| **Sub-epidemic II** | 5 | $\geq 1$ (*n*) | Yes | Yes | Yes |

The 2-parameter logistic growth model is useful as a simple benchmark for comparing the performance of the more complex models. The well-known 3-parameter Richards model extends the logistic growth model to include an additional parameter to allow for asymmetry in the decline of the epidemic curve (45, 46). If the data follow a symmetric logistic trajectory, then the logistic model can accurately fit the data. However, if the incidence is asymmetric, then the Richards model will yield a better fit.

We also introduce and apply an extension of the Richards model that consists of 5 parameters and an endemic state; therefore, we denote this the “endemic Richards” model (33). If the epidemic declines to a steady state or endemic level, rather than declining to extinction, the Richards model will under-predict the future incidence. When this happens, short-term forecasts derived using the endemic Richards model tend to outperform the simple Richards.

The model is then extended to a 6 parameter “double logistic” model that supports trajectories with double peaks or temporary steady states followed by a secondary decline (Figure S2) (33). When data points fall outside the PI of the endemic state assumed by the endemic Richards model, a decline greater than the assumed level of statistical noise is indicated; meaning, it is likely a true decline, rather than stochasticity. Therefore, the endemic Richards model will overpredict the incidence, and the double logistic model will be more appropriate.

We introduce a multi-model approach (see next section) that sequentially uses the four single-equation models mentioned above. For this purpose, we compare the models in order of increasing complexity (Table 1) and assess prediction interval coverage to determine which model to employ for the forecast. Our multi-model algorithm is summarized in Figure 1.

**Figure 1.**
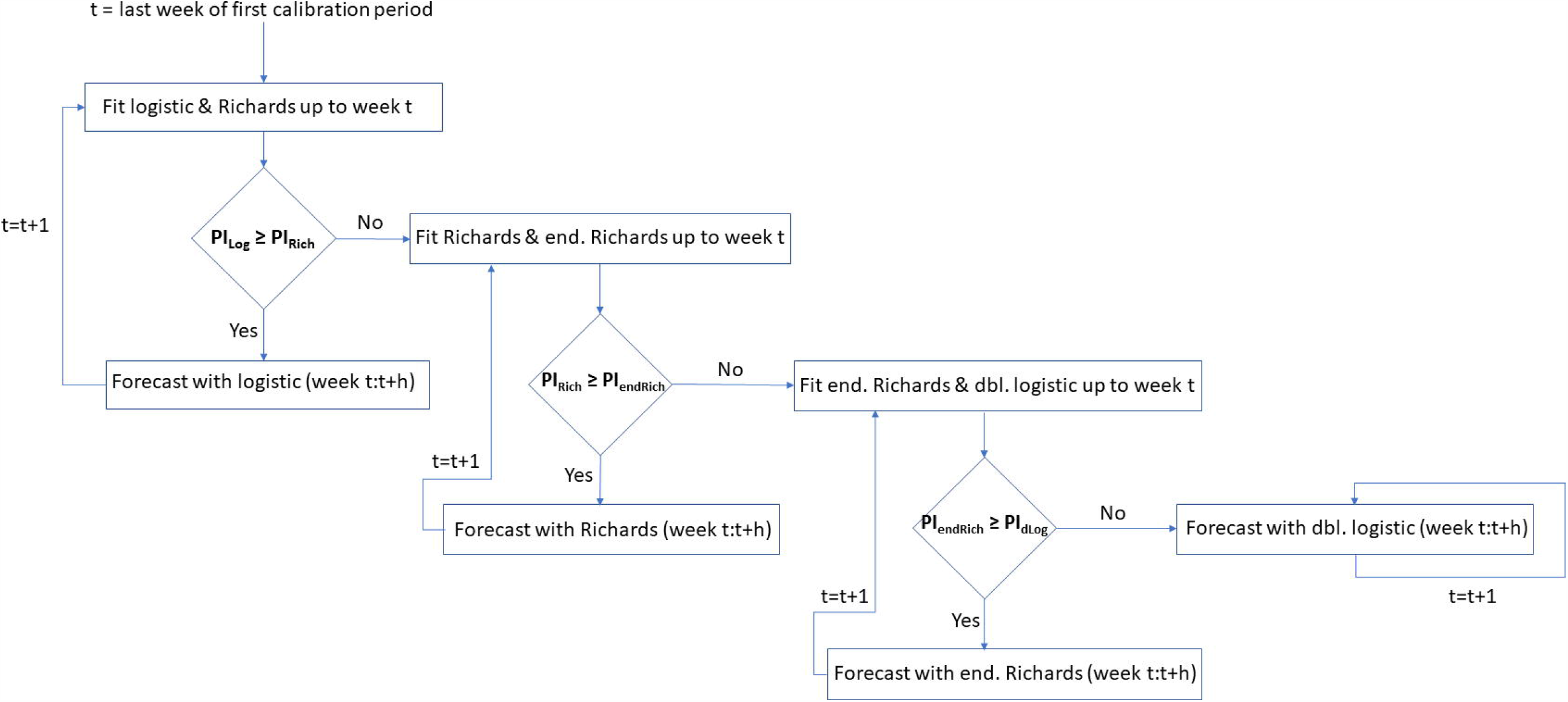
Schematic for the performance-based multi-model approach. This describes the process of choosing models to provide forecasts for each weekly projection.

The most flexible model we use is a sub-epidemic wave model that supports complex temporal dynamic patterns, such as oscillating dynamics leading to endemic states or damped oscillations (30). We incorporate two variations of sub-epidemic decline: exponential decline, as presented in (30), and a new extension with an inverse decline function; each of the variations includes 5 fitting parameters. This approach assumes that multiple underlying sub-epidemics shape the aggregate reported epidemic curve, where each sub-epidemic is modeled using a generalized logistic growth model. These combine to create an epidemic wave composed of *n* overlapping sub-epidemics modeled using a system of *n* coupled differential equations.

### Multi-model approach

For the multi-model approach, we compare the four single-equation models in order of increasing complexity (Table 1), and we assess the PI coverage of the calibration period to determine when/if to switch models, as summarized in our schematic shown in Figure 1.

We begin at the initial forecasting week by comparing the calibration PI coverage between the logistic and Richards models. When the calibration PI coverage of the logistic model is greater than or equal to the PI coverage of the Richards model, we provide forecasts with the logistic model. When the PI coverage of the Richards is greater, we then switch to comparing the Richards to the endemic Richards, and the iterative process continues as such (Figure 1).

We define the *forecasting phases* as the time intervals corresponding to the Situation Reports for which each model is used. That is, each time the method switches to a new forecasting model, a new forecasting phase is initiated, and there will be as many forecasting phases as models used. Notably, any number of the 4 models could make up the multi-model approach. For example, if the Richards model provides higher PI coverage than the logistic model at time *t*_*switch*_ and the endemic Richards model has higher PI coverage than the Richards at time *t*_*switch*_, then the Richards model would not be used for any forecasts (Figure 1). The models are analyzed in the explicit order reported, and once a model is switched to, there is no switching back to simpler models.

## Results

We compare the calibration and real-time short-term forecasting performance of the seven models in Table 1 on the major Ebola resurgence between March 11, 2019 and September 23, 2019. We further assess performance within each forecasting phase, as defined by the multi-model approach. The Supplement contains additional figures of the model fits (Figures S3 – S8).

### Forecasting phases

As explained in the methods, we define our *forecasting phases* by assessing the calibration PI coverage of the four single-equation models as defined by the multi-model approach (Figure S3). The following forecasting phases were obtained: weekly forecasts with the logistic model for March 11 – April 1, 2019 (data from Situation Reports 33 – 36), with the Richards model for April 8 – June 10, 2019 (Situation Reports 37 – 46), with the endemic Richards model for June 17 – July 22, 2019 (Situation Reports 47 – 52), and with the double logistic for July 29 – September 23, 2019 (Situation Reports 53 – 61) (Figure 2). We will refer to these consecutive forecasting phases as: Incline, Oscillating I, Oscillating II, and Decline, respectively. These break points based on PI coverage are also consistent with the timing of where the models begin to deviate with respect to each of the other calibration performance metrics - MSE, MAE, and MIS (Figure 3).

**Figure 2.**
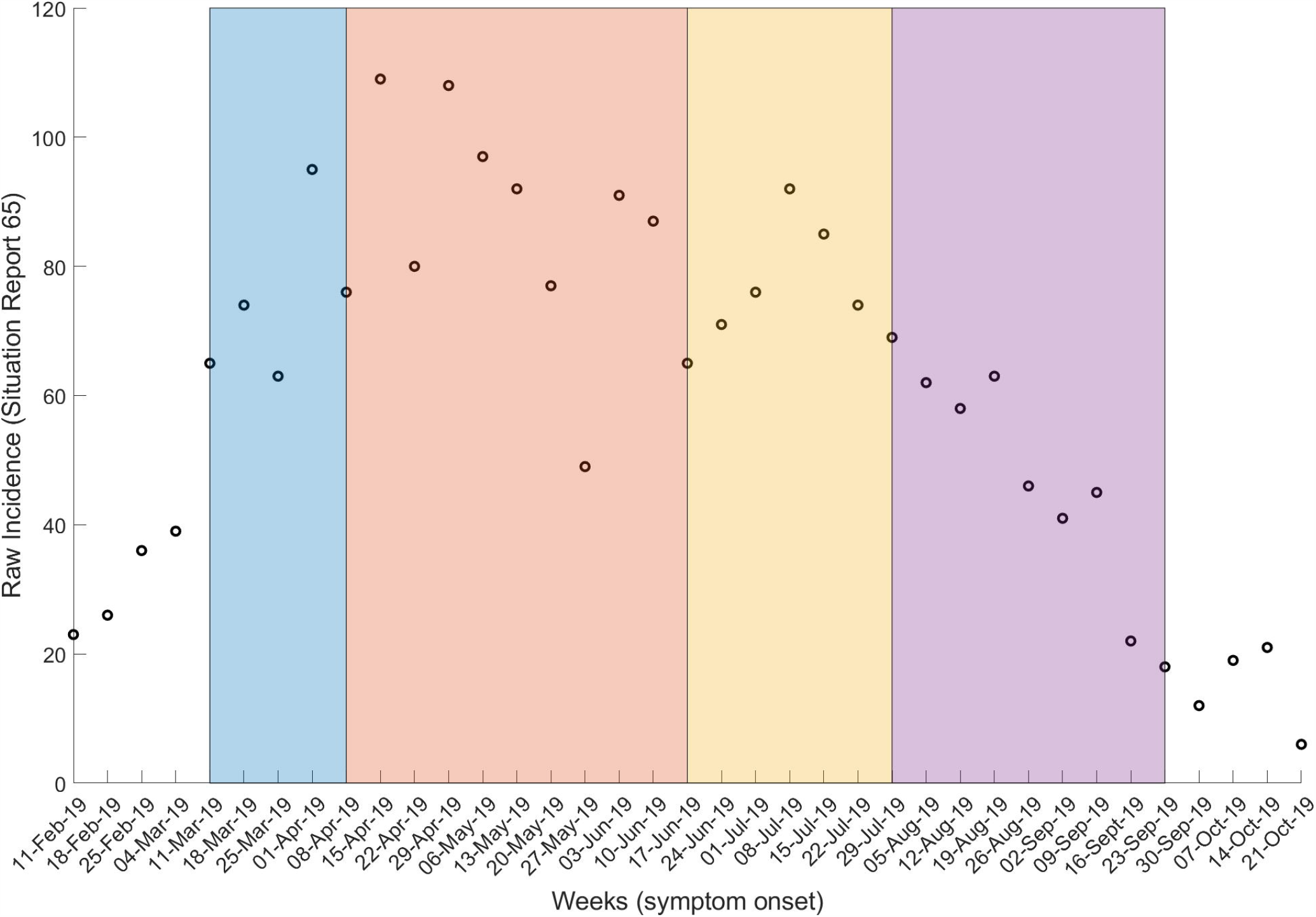
Visual representation of the forecasting phases defined by the multi-model approach. Raw data from Situation Report 65 is shown with the periods for which each model was used: logistic growth model (blue), Richards model (red), endemic Richards (yellow), and double logistic (purple).

**Figure 3.**
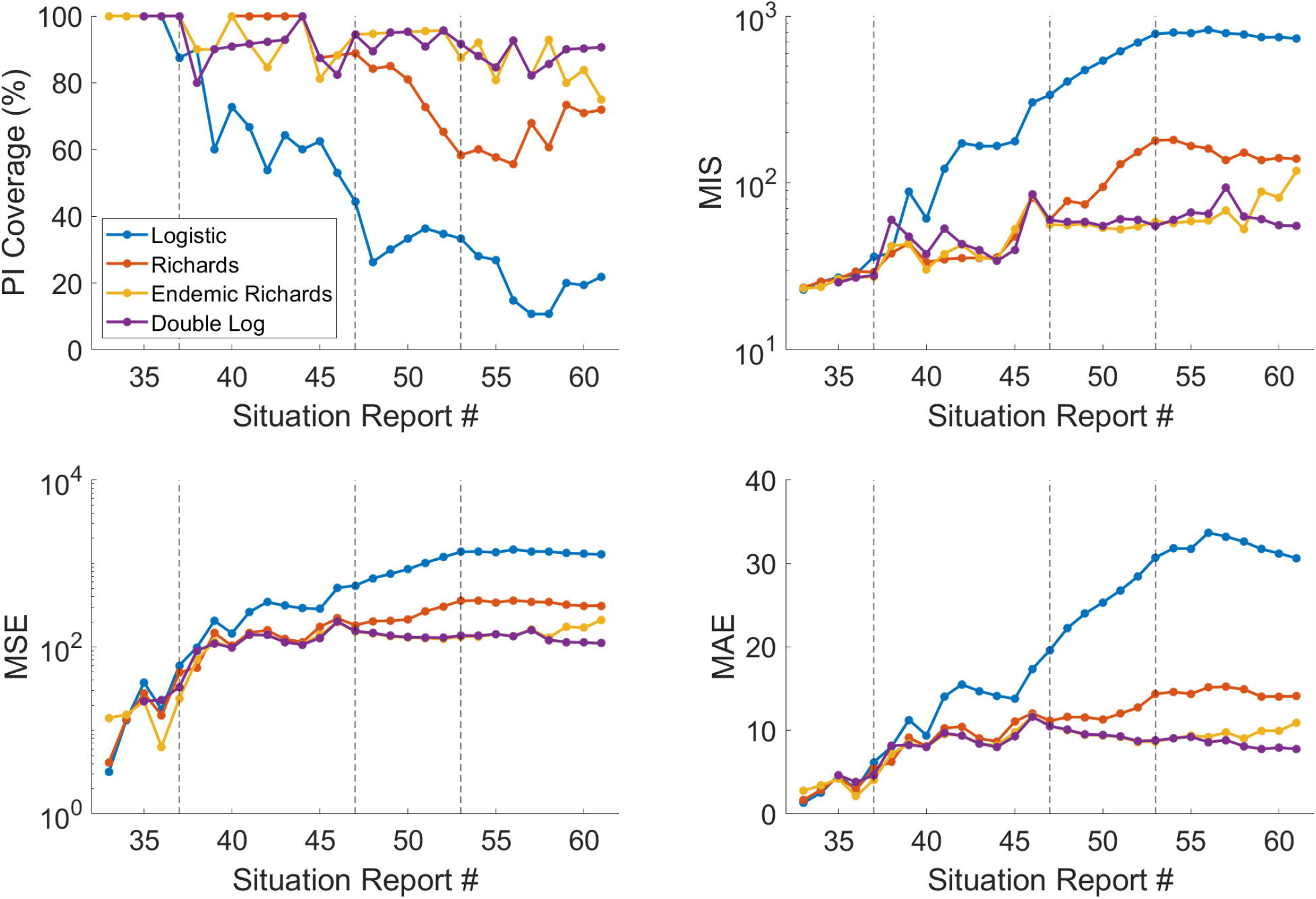
Calibration period metrics for the 4 models included in the performance-based multi-model approach: logistic growth model (blue), Richards model (red), endemic Richards (yellow), and double logistic (purple). Prediction interval coverage is used to indicate the ‘switch’ in models

In general, the resulting forecasting phases obtained by our multi-model approach are consistent with our rationale for incorporating the dynamic models supporting different dynamics. Data from February 11 – April 1 represent the early growth dynamics of the 2019 resurgence; thus, we define the first phase (March 11 – April 1) as the incline phase, for which the simple logistic model is sufficient for fitting the data (Figure S3).

In the next phase beginning April 8, Oscillating I, the outbreak begins to fluctuate, and the Richards model outperforms the logistic model. As new observations are added to the weekly incidence curve, the deviations between the logistic model and data become more pronounced (Figure 3 & S3). The trajectory continues on a sustained oscillating pattern through the next phase, Oscillating II, so the endemic Richards model provides a better model fit than the simple Richards model.

On July 29, the switch to the final model and the Decline phase is initiated (Figure 3). From July 29 – September 2 (Situation Reports 53 – 58), the endemic Richards and double logistic model perform comparably in each of the calibration metrics. However, the double logistic model outperforms the endemic Richards model between September 9 – September 23 (Situation Reports 59 – 61). This is the point where the trajectory falls outside the 95% prediction interval obtained using the endemic Richards model, suggesting a need for a model that can capture the declining trend (Figures S6 & S7). The double logistic model outperforms the other single-equation models in capturing the full national incidence pattern up to September 23 (Table 2, Figure 3).

**Table 2.**
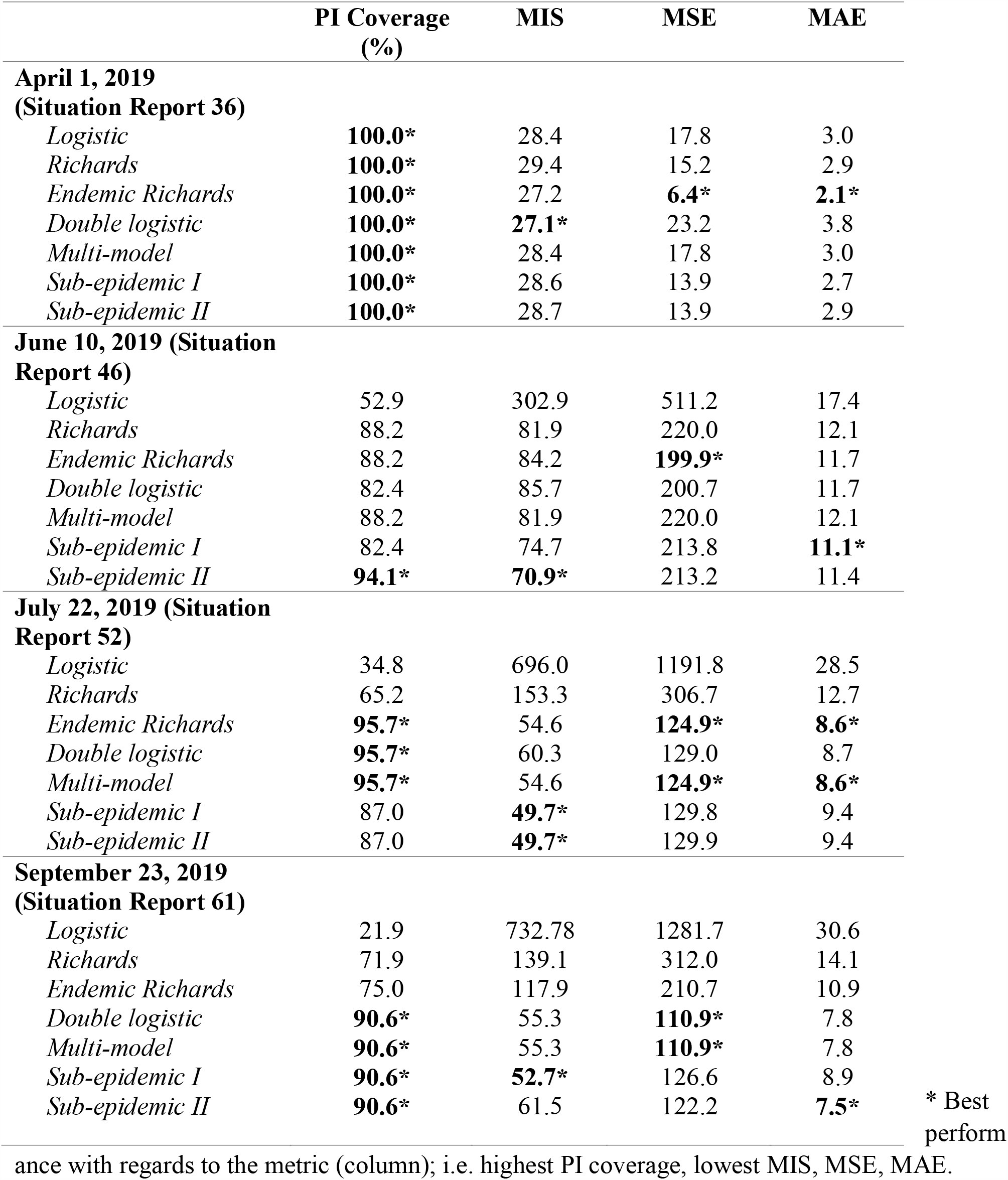
Calibration performance of each model for the last calibration endpoint of the time intervals defined by the multi-model approach

### Calibration performance

The calibration performance metrics across phases, based on the last date of calibration within each of the four phases, are given in Table 2. For data through the Incline phase, each of the models provides 100% PI coverage and very similar MIS, with the double logistic having the lowest (MIS=27.1), followed by the endemic Richards (MIS=27.2). The endemic Richards model has significantly lower MSE and MAE for the Incline phase compared to the other models (Table 2).

For data through Oscillating I, the sub-epidemic model type II has the highest PI coverage (94.1%) and lowest MIS (70.9), while the endemic Richards and sub-epidemic type I have the lowest MSE and MAE, respectively (Table 2). For data through Oscillating II, the endemic Richards, multi-model, and the double logistic model have the highest PI coverage (95.7%), while sub-epidemic types I and II have the lowest MIS (49.7). The endemic Richards, which corresponds with the multi-model approach for Oscillating II, also has the lowest MSE and MAE.

When fitting all the available data through September 23, or the Decline phase, the double logistic, multi-model, and both sub-epidemic models perform best in terms of PI coverage (90.6%); however, the other metrics are split between these models (Table 2). The three simplest models perform poorly on the full data, supporting the need for more flexible models to capture the complex dynamics of the epidemic.

Weekly calibration performance across the entire incidence curve using the double logistic model, the performance-based multi-model approach, and the two sub-epidemic models are displayed in Figure 4. Goodness-of-fit metrics do not point to a single winner or ‘best’ model (Figure 4). In terms of mean model fit and error, the models perform comparably with regards to MSE and MAE. The models show variation in PI coverage and MIS; however, the curves repeatedly overlap, suggesting that there is not necessarily a clear best model across the full epidemic trajectory.

**Figure 4.**
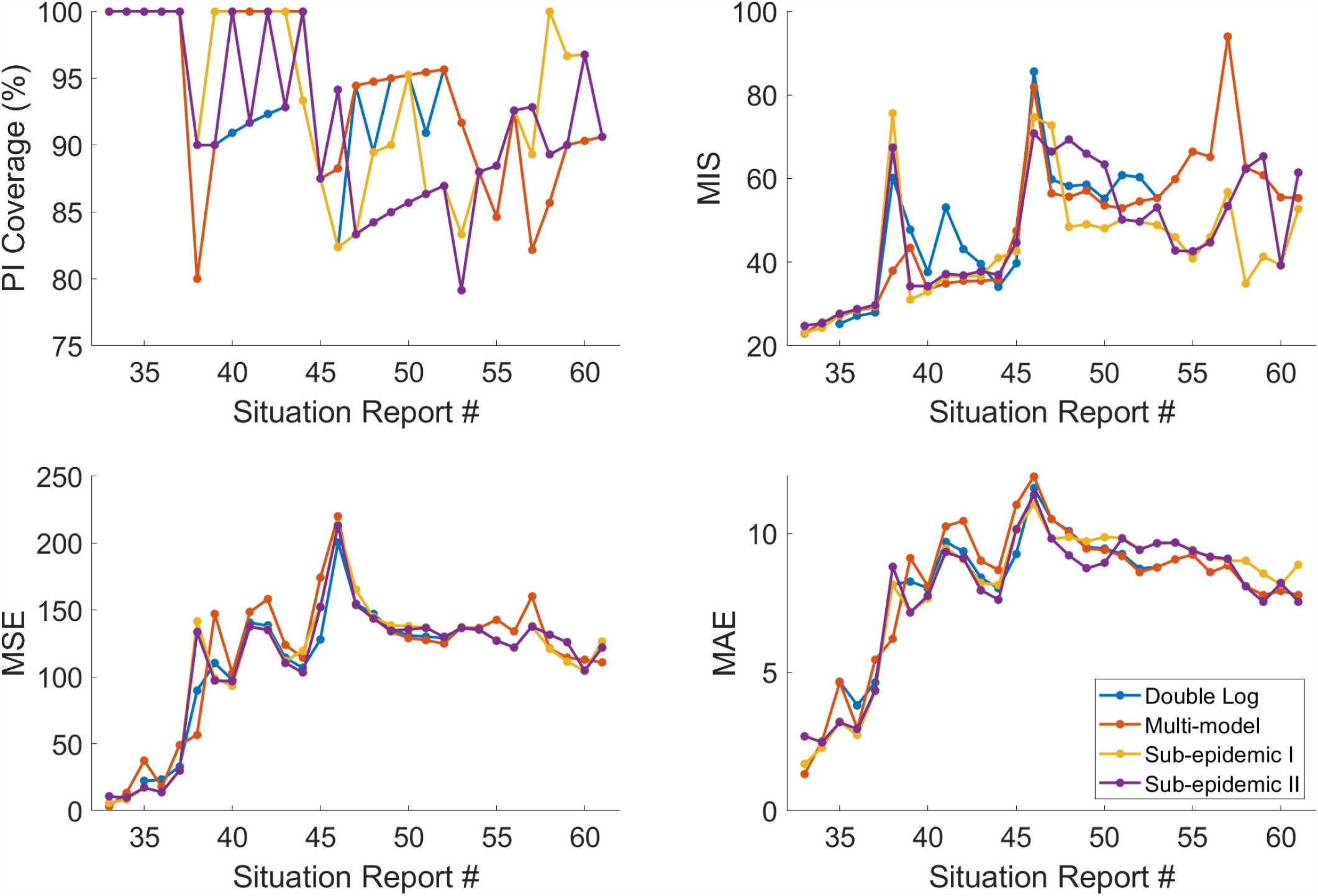
Calibration period metrics across all Situation Reports for the double logistic (blue), multi-model (red), and sub-epidemic types I (yellow) and II (purple).

**Figure 5.**
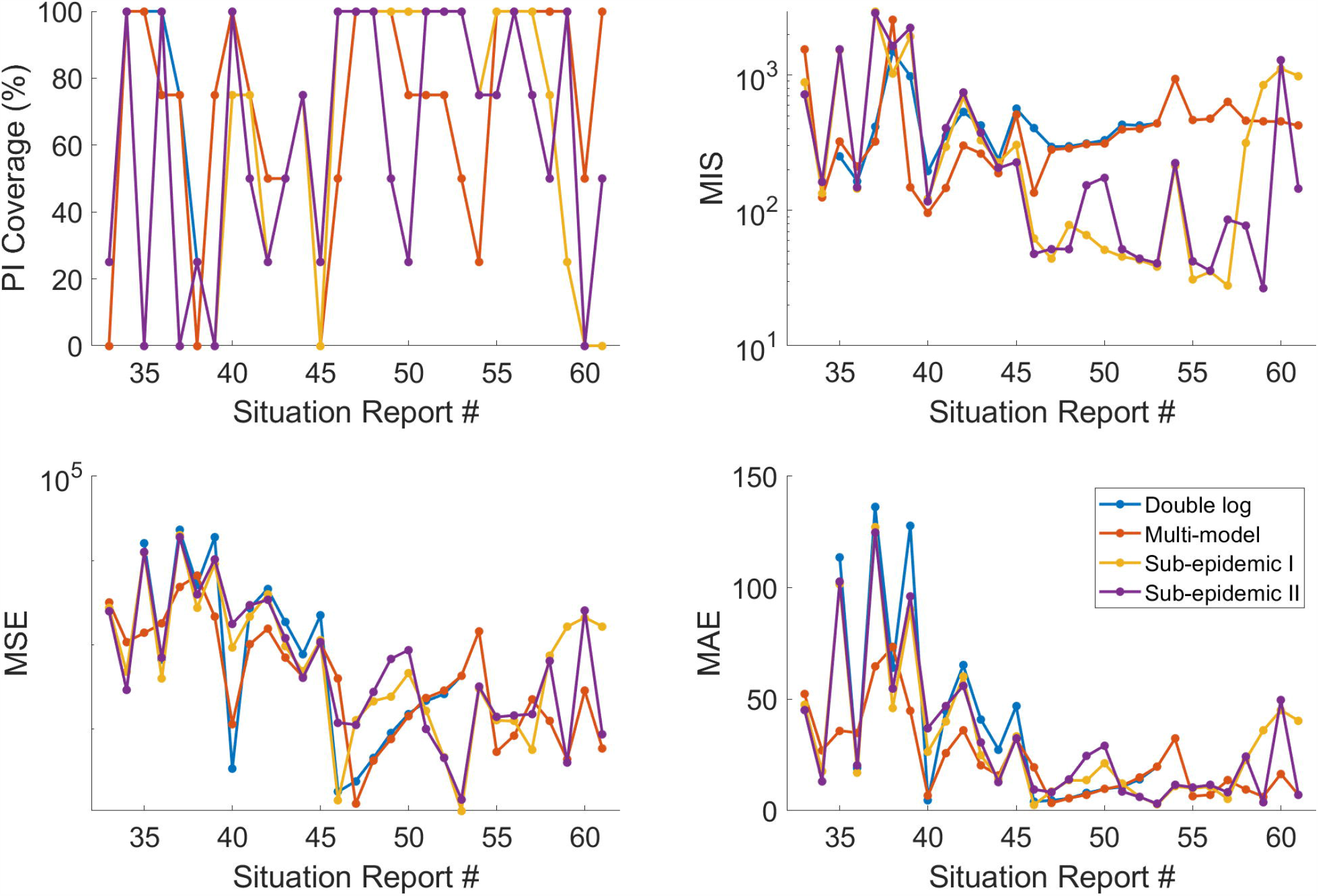
Forecasting period metrics for *h* = 4 across all Situation Reports for the double logistic (blue), multi-model (red), and sub-epidemic types I (yellow) and II (purple).

### Forecasting performance

The forecasting results by phase are presented in Table 3. For the Incline phase, the endemic Richards model provides forecasts with substantially lower MSE and MAE than any other model. The double logistic model has the highest PI coverage (100%) and lowest MIS (207.3); however, the MSE is more than 14 times higher than that of the endemic Richards (Table 3). Thus, the high coverage can be attributed to very wide prediction intervals (Figure S6).

**Table 3.**
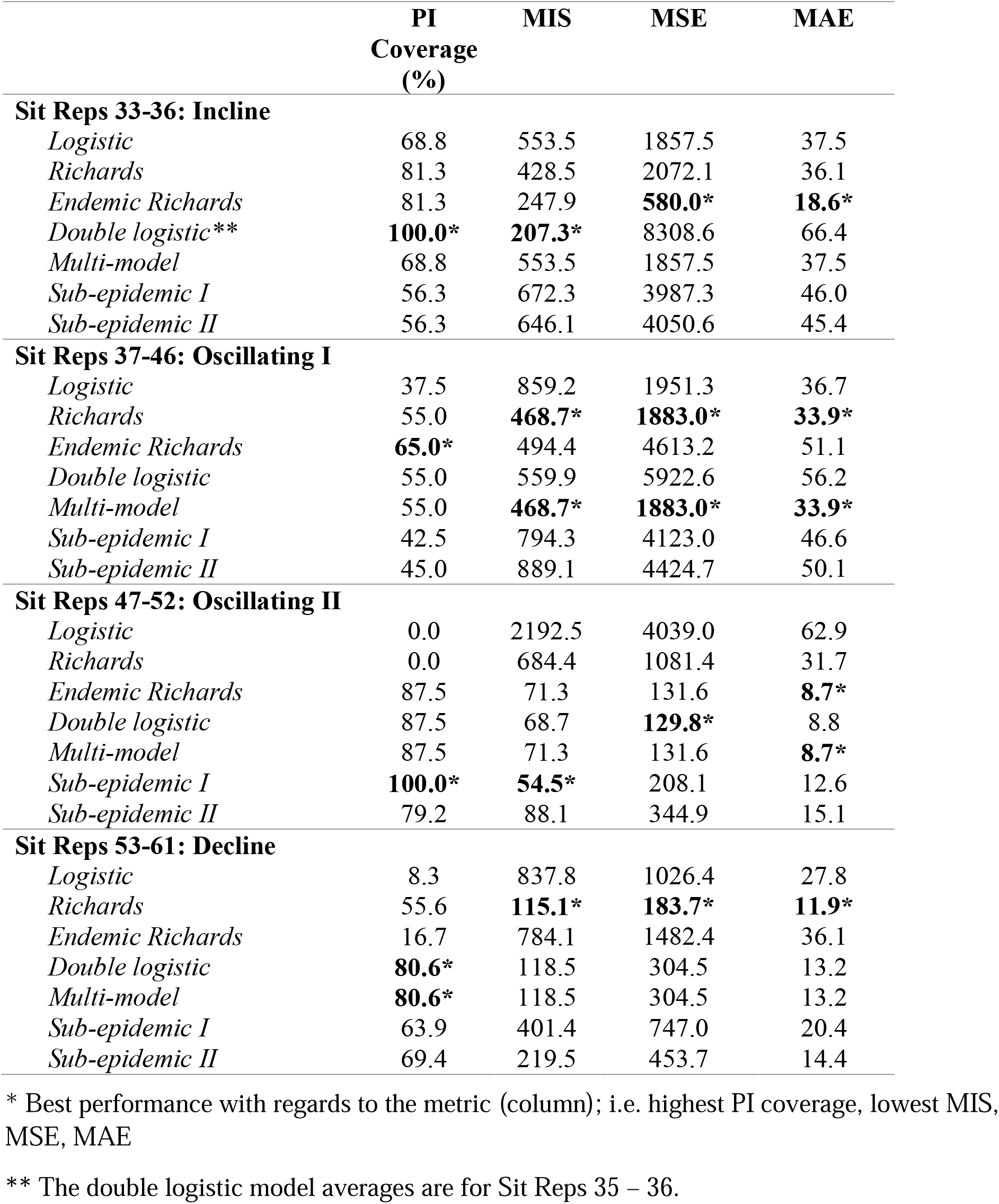
Average forecasting performance of 4-week ahead forecasts across Situation Reports within four distinct forecasting phases, as defined by the multi-model calibration results (corresponding with Figure 2). Note: the metrics for the multi-model approach will be equivalent to the individual model used in their respective time intervals.

For Oscillating I, the endemic Richards model provides the highest PI coverage of future data points (65.0%), while the Richards model has the lowest MSE, MAE, and MIS, which correlates with the multi-model approach having the lowest error and MIS as well (Table 3).

As more complicated dynamics emerge, the simpler models fail to predict the epidemic trajectory accurately. For Oscillating II, both the logistic and Richards models have PI coverage of 0% with high error (Table 3). The sub-epidemic model type I, outperforms all other models on PI coverage (100%) and MIS (54.5) for this phase, indicating it has high PI coverage without significantly higher error. The endemic Richards, double logistic, and multi-model approach yield the lowest MAE and MSE for Oscillating II.

For the final phase, the Decline phase, the double logistic and multi-model approach yield the highest forecast PI coverage (80.6%). Interestingly, the simple Richards model provides forecasts with the lowest MIS, MSE, and MAE for the last phase; however, PI coverage is only 55.6% (Table 4). Further, if we had continued conducting forecasts past the end of the study period, the Richards model would have failed to capture the continued endemic state observed. The double logistic and multi-model approach rank second in MIS, MSE, and MAE, so while there is not a clear best model, the double logistic and multi-model approach highly perform across all of the metrics.

## Discussion

We conducted a systematic comparison of seven models for short-term real-time forecasting the ongoing 2018-20 Ebola outbreak in the DRC. A well-defined performance-based approach was used to identify distinct epidemic phases for which to employ different models to capture the complex trajectory of the epidemic. By using different models for different phases of an epidemic, the approach can account for significant changes in transmission dynamics over the course of the outbreak, ranging from a simple logistic curve to incidence curves with oscillatory behavior, as observed in the DRC (Figure 3).

The first defined phase, Incline, covers the sharp increase in cases observed in late February – early March, which followed an increase in armed attacks, including the burning of Ebola treatment centers in Katwa and Butembo (31). Specifically, February 2019 recorded the highest monthly incidence of armed attacks, corresponding with the increase in cases observed in the Incline phase. The following two phases represent oscillating dynamics, which correspond with continued violent attacks and increasing community resistance that deterred response activities (28, 31). As the incidence of violent attacks decreased in July 2019, cases leveled out and eventually showed a substantial decline for the final Decline phase.

The double logistic model and the sub-epidemic models (types I and II) provide the best fit to the incidence trajectory through the study period (Table 2); however, in general, goodness-of-fit was not found to be correlated with forecasting performance. While the sub-epidemic models often provide the best fit to the calibration data (Table 2), they were less successful in forecasting short-term dynamics of the epidemic (Table 3). We observed that the sub-epidemic forecasts in the decline phase perform poorly, as the trajectory is declining while the models are predicting another upturn in cases or sub-epidemic waves (Figure S7 & S8). The sub-epidemic model, with an inverse decline function (type II), is more successful at capturing the future declining trajectory in the Situation Reports 58 – 61, whereas the version with the exponential decline (type I) cannot predict the declining trend observed in following weeks (Table 3; Figure S7 & S8).

The multi-model approach provides the most consistent forecasts, in terms of average MSE and MAE, throughout our study period (Table S2). Even when broken into phases, the multi-model approach performs best in at least one of the forecasting metrics for each forecasting phase, which was not the case for any other model (Table 3). This general multi-model approach can be adapted to other epidemic scenarios, such as epidemics of emerging pathogens or those occurring in regions with unstable sociopolitical climate, as the models are phenomenological and do not require biological information or knowledge of specific disease transmission processes. However, the four models incorporated here may not be appropriate for all outbreak scenarios. For example, these models do not allow for a higher second peak. This approach would also have failed to predict the February 2019 resurgence, like the other early projections.

The general multi-model approach can be adapted to incorporate any sequence of models. For disease outbreaks with more epidemiological data, specific disease mechanisms can be incorporated in compartmental models that increase in complexity as more outbreak characteristics are elucidated. As model complexity increases, however, the uncertainty of model estimates must be considered. Here, the models build upon each other and have very similar estimates for the early phase, so we rely on PI coverage as our ‘switch’ metric to remain at a simpler model while they all have equivalent coverage. This could potentially be problematic for more complex models, as very wide intervals, such as (0, inf.), would perform ‘better’ in terms of PI coverage, leading to high uncertainty in forecasts. In this situation, one may consider MIS to classify the phases, rather than PI coverage.

Another modeling approach rapidly gaining traction in epidemiological literature is ensemble modeling, which involves incorporating multiple models in a complementary manner (47-49). Rather than a sequential multi-model approach, future work could rely on an ensemble modeling approach based on a combination of simple dynamic models. With an ensemble approach, we would have the option to base the contribution of each model on calibration performance, rather than choose one model based on calibration as we did here. Another option is to weight the models based on the forecasting performance of prior weeks; however, in this study, forecasting performance in one phase is not clearly predictive of performance in the following phase (Table 3). The use of an algorithm like that presented here could supplement ensemble models to define distinct epidemic phases, which may yield better projections compared to separating data by standard intervals.

Reviews of real-time forecasting throughout the historic 2014-15 Ebola epidemic found that forecasting uncertainty is higher in the beginning stages of an outbreak and decreases over time (16, 17); however, this was not observed in the 2018-20 Ebola epidemic. Fluctuations in error and MIS do not reveal a declining pattern in forecast uncertainty as the epidemic progresses. This highlights the challenge of forecasting the complicated dynamics of this epidemic, where increasing the amount of available data does not necessarily decrease the uncertainty of estimates.

The unpredictable social components of the epidemic on the ground in the DRC are major limitations to the study. While we adjust the reported data in real-time by the week of symptom onset, we do not know the true stable incidence pattern when forecasts are generated. Further, we rely on phenomenological models, which are particularly valuable for providing rapid predictions of epidemic trajectory in complex scenarios; however, they do not incorporate disease-specific mechanisms or explicitly account for interventions. The sub-epidemic modeling framework is the most flexible presented here, and only the two sub-epidemic models can predict a resurgence. For the variations applied here, the models cannot predict a larger second wave, as observed in the DRC outbreak. Therefore, none of the models employed here would have anticipated the 2019 disease resurgence, but, when applied from the start of the resurgence, they can be used to forecast the following trajectory in real-time.

In conclusion, while the forecasting models introduced here are relatively simple, we are encouraged by the short-term forecasting performance of the model extensions, especially when applied to such a complex, non-traditional epidemic trajectory. This work suggests a multi-model framework, such as the one presented here, can identify distinct forecasting phases that allow the model to adjust for changing dynamics. Further, the general approach is flexible and can be adapted to many different model combinations and outbreak scenarios. Forecasting challenges during the DRC outbreak underscore the need for more research into flexible real-time forecasting approaches, especially when the dynamics exhibit complex temporal patterns.

## Data Availability

Data will be made available in an online repository upon acceptance of the manuscript.

## Acknowledgements

We thank Homma Rafi (Director of Communications, School of Public Health, Georgia State University) for creating and maintaining the online record of weekly short-term forecasts.

## Funding

GC is supported by NSF grants 1610429 and 1633381 and NIH R01 GM 130900

## Ethics

Not applicable.

## Data, code, and materials

Data will be made available in an online repository upon acceptance of the manuscript.

## Competing Interests

Not applicable.

## Author Contributions

KR and GC conducted the forecasts and data analysis; AT retrieved and adjusted data; KR wrote the first draft of manuscript; all authors contributed to writing and revising subsequent versions of the manuscript. All authors read and approved the final manuscript.

## Notes

### Competing Interest Statement

The authors have declared no competing interest.

